# Nontuberculous Mycobacteria Species in Patients with Concurrent COVID-19 and Mycobacterial Disease

**DOI:** 10.1101/2025.10.02.25337223

**Authors:** Platon Eliseev, Alexandra Bairakovа, Anastasia Kazyulina, Evgeny Kuznetsov, Anastasia Samoilova, Irina Vasilieva

## Abstract

**Background:** The incidence of nontuberculous mycobacterial (NTM) diseases is increasing globally, with risk factors remaining poorly understood. COVID-19 may alter respiratory tract conditions affecting NTM colonization and infection patterns.

**Objective:** To evaluate species diversity of NTM isolated from patients with NTM-associated diseases with and without a history of COVID-19.

**Methods:** We analyzed 464 clinical NTM isolates from Russian patients stratified by COVID-19 status (COVID-19+, n=271; COVID-19−, n=193). Diagnosis verification followed ATS/ERS/ESCMID/IDSA Clinical Practice Guidelines. Species identification employed MALDI-TOF mass spectrometry and molecular techniques. Statistical analyses used chi-square and Fisher’s exact tests with 95% confidence intervals.

**Results:** Slowly growing species predominated in both groups (97.9% vs 98.2%, p>0.05). COVID-19 was associated with a significant 28% reduction in Mycobacterium avium complex (MAC) prevalence (94.8% vs 66.8%, p<0.05), primarily due to decreased M. avium isolation (80.3% vs 52.0%, p<0.05). Conversely, rare NTM species showed increased prevalence in COVID-19+ patients: M. kansasii complex (0% vs 8.1%, p<0.05), M. terrae complex (0% vs 3.3%, p<0.05), M. simiae complex (0.5% vs 7.0%, p<0.05), and unclassifiable species (2.6% vs 12.9%, p<0.05).

**Conclusions:** COVID-19 significantly alters the NTM species spectrum in patients with NTM-associated diseases, with reduced MAC prevalence and increased detection of rare NTM species. These findings highlight the importance of species-level identification and may require adaptation of diagnostic and therapeutic approaches in post-COVID patients.

## Introduction

Nontuberculous mycobacteria (NTM) are opportunistic environmental pathogens of increasing clinical significance worldwide. The genus Mycobacterium comprises various species of mycobacteria, including NTM – a diverse group of opportunistic microorganisms found in the environment that were initially not considered pathogenic to humans. Currently, the incidence of mycobacterioses (diseases caused by NTM) is increasing worldwide [6,13,16], however, the underlying causes of this trend remain unclear [0]. The risk factors include the presence of comorbidities [6,7] and various conditions involving obstructive, restrictive, or mixed pulmonary function disorders [9]. The primary source of NTM infection in humans is environmental exposure to contaminated water, soil and biofilms [2,12].

Clinical manifestations vary considerably; some patients show NTM colonization without disease requiring treatment [3], while others develop clinically significant mycobacterial disease requiring prolonged antimicrobial therapy [4]. The disparities in clinical presentation and its interpretation, the absence of mandatory reporting for NTM-associated diseases, and the consequent lack of official statistics complicate epidemiological assessment and surveillance efforts. The mechanisms underlying the pathogenesis of nontuberculous mycobacterial diseases remain incompletely understood and warrant additional research [5,8,21,22]. The etiological structure of NTM-associated diseases is diverse and can depend on various factors. The presence of NTM is often accompanied by other infectious diseases, and such coinfections can arise due to immune response impairments, which may be both a cause and a consequence of the comorbid pathology. [9,10]. The COVID-19 pandemic has raised concerns about altered susceptibilities to concurrent infections. Global studies suggest COVID-19 may influence the progression of co-occurring infections [11,23,24], including potential interactions with NTM [9,20]. COVID-19 and NTM infections share immunopathological features, including T-cell activation and interferon signaling pathway induction [25,26,27]. In COVID-19, an antiviral response develops aimed at eliminating the pathogen, whereas in NTM infections, a prolonged cell-mediated immune response is formed, providing control over the persistent bacterial infection. It is important to note that inflammatory processes and the resulting immune imbalance in these diseases can mutually potentiate, creating prerequisites for a more severe course of coinfection. Of particular concern is the ability of COVID-19 to cause transient immunosuppression and modulation of the immune response, which may adversely affect the control of NTM infection, increasing the risk of its progression and complications [20]. Thus, the identification of clinically relevant NTM pathogens causing NTM-associated diseases in post-COVID-19 individuals is of particular contemporary importance.

Despite these concerns, comprehensive data on COVID-19 and NTM interactions remain limited, particularly regarding species-specific changes in NTM epidemiology. This study aimed to evaluate the species diversity of NTM isolated from patients with NTM-associated diseases with and without documented COVID-19 infection in the Russian Federation.

## Materials and Methods

### Study Design and Population

This retrospective cross-sectional study analyzed 464 clinical NTM isolates obtained from patients diagnosed with NTM-associated diseases across various regions of the Russian Federation. Data on NTM isolates as well as past history of COVID-19 was accessed from laboratory register on 1st of October 2024. Authors didn’t’ have access to information that could identify individual participants during or after data collection. Based on data isolates were further stratified into two groups: those with documented SARS-CoV-2 exposure (COVID-19+, n=271) and those without evidence of infection (COVID-19−, n=193).

### Diagnostic Criteria

Diagnosis of NTM-associated diseases was conducted in accordance with the clinical guidelines Treatment of Nontuberculous Mycobacterial Pulmonary Disease: An Official ATS/ERS/ESCMID/IDSA Clinical Practice Guideline [15], what ensured a unified methodological approach to diagnosis verification across all healthcare institutions. COVID-19 was defined according to the interim national clinical guidelines.

### Laboratory Methods

Methods of laboratory diagnostics included standard procedures for the isolation and differentiation of NTM [14]. Species-level identification employed matrix-assisted laser desorption/ionization time-of-flight (MALDI-TOF) mass spectrometry using a Bruker Daltonics instrument (Germany). Isolates were classified by growth characteristics (slow vs rapid growers) and assigned to recognized phylogenetic complexes using molecular and phenotypic criteria.

### Statistical Analysis

Data analysis utilized Microsoft Excel, Biostatistics for Windows, and MedCalc software. Intergroup comparisons employed chi-square tests (for homogeneity and Pearson’s), Fisher’s exact tests, and 95% confidence intervals for species prevalence estimates. Statistical significance was defined as p<0.05.

## Results and discussion

Initially, NTM isolates were stratified based on their growth kinetics (rapid vs. slow growers) and subsequently assigned to recognized phylogenetic complexes using molecular and phenotypic criteria. – *M. avium complex, M. fortuitum complex, M. terrae complex, M. abscessus complex* and other groups [17,18]. On the one hand, this classification simplifies the understanding of the overall prevalence patterns; on the other hand, it introduces new challenges, which involve identifying and assessing the clinical significance of individual species, including selecting drug therapy regimens based on drug resistance profiles within specific complexes.

Analysis revealed that slowly growing NTM predominated in both groups (COVID-19−: 97.9%, 95% CI: 95.7-99.0; COVID-19+: 98.2%, 95% CI: 95.9-99.3; p>0.05). Rapidly growing NTM were rare in both cohorts (COVID-19−: 2.1%, 95% CI: 0.8-5.1; COVID-19+: 1.8%, 95% CI: 0.7-4.2; p>0.05). Overall, these findings are consistent with studies by other authors, who also report a higher frequency and prevalence of slowly growing NTM species [18].

The species diversity of isolates obtained from patients with NTM-associated diseases allowed for the classification of the detected isolates into five distinct complexes based on growth characteristics. Among slow growers following NTM were obsereved: *Mycobacterium avium complex* (*M. avium, M. intracellulare, M. chimaera, M. colombiense. M. celatum*), *Mycobacterium terra complex (M. heckeshornense), Mycobacterium kansasii complex (M. gastri), Mycobacterium simiae complex (M. lentiflavum, M. asiaticum)*, as well as species not assigned to established NTM complexes – *M. malmoense, M. europaeum, M. branderi* и *M. xenopi*. Rapidly growing isolates were further categorized into recognized complexes, such as *M. abscessus* complex (*M. abscessus subspecies abscessus, M. abscessus subspecies massiliense*), *M. fortuitum* complex (*M. fortuitum*) and unclassifiable species *M. stephanolepidis* (taбл.2).

**Table 2.** Distribution of slowly and rapidly growing NTM species isolated from patients with NTM-diseases with or without prior history of COVID-19.

|  |  | COVID-19– |  |  | COVID-19+ |  |  |  |  |
| --- | --- | --- | --- | --- | --- | --- | --- | --- | --- |
|  | NTM species | n=193 | % | 95% CI | N =271 | % | 95% CI | Difference | p<0.05 |
| **I** | **Slow-growers** | 189 | 97,9% | 95,7 – 99,0 | 266 | 98,2% | 95.9 – 99.3 | 0,2% | no |
| 1 | Mycobacterium avium complex: | 183 | 94,8% | 91,9 – 96,7 | 181 | 66,8% | 61.1 – 72.1 | -28,0% | yes |
|  | *M. avium* | 155 | 80,3% | 74,9 – 84,8 | 141 | 52,0% | 45.9 – 58.1 | -28,3% | yes |
|  | *M. chimaera/ intracellulare* | 24 | 12,4% | 8,4 – 17,8 | 23 | 8,5% | 5.6 – 12.5 | -3,9% | no |
|  | *M. colombiense* | 1 | 0,5% | 0,1 – 1,7 | 9 | 3,3% | 1.7 – 6.2 | 2,8% | yes |
|  | *M. celatum* | 3 | 1,6% | 0,5 – 3,7 | 8 | 3,0% | 1.4 – 5.7 | 1,4% | no |
| 2 | Mycobacterium kansasii complex: | 0 | 0,0% | 0,0 – 1,9 | 22 | 8,1% | 5.2 – 12.3 | 8,1% | yes |
|  | *M. kansasii* | 0 | 0,0% | 0,0 – 1,9 | 13 | 4,8% | 2.7 – 8.1 | 4,8% | yes |
|  | *M. gastri* | 0 | 0,0% | 0,0 – 1,9 | 9 | 3,3% | 1.7 – 6.2 | 3,3% | yes |
| 3 | Mycobacterium terra complex: | 0 | 0,0% | 0,0 – 1,9 | 9 | 3,3% | 1.7 – 6.2 | 3,3% | yes |
|  | *M. heckeshornense* | 0 | 0,0% | 0,0 – 1,9 | 9 | 3,3% | 1.7 – 6.2 | 3,3% | yes |
| 4 | Mycobacterium simiae complex: | 1 | 0,5% | 0,1 – 1,7 | 19 | 7,0% | 4.5 – 10.7 | 6,5% | yes |
|  | *M. lentiflavum* | 1 | 0,5% | 0,1 – 1,7 | 11 | 4,1% | 2.2 – 7.3 | 3,5% | yes |
|  | *M. asiaticum* | 0 | 0,0% | 0,0 – 1,9 | 8 | 3,0% | 1.4 – 5.7 | 3,0% | yes |
| 5 | unclassifiable species: | 5 | 2,6% | 1,0 – 5,8 | 35 | 12,9% | 9.2 – 17.8 | 10,3% | yes |
|  | *M. xenopi* | 3 | 1,6% | 0,5 – 3,7 | 10 | 3,7% | 1.9 – 6.8 | 2,1% | no |
|  | *M. malmoense* | 2 | 1,0% | 0,2 – 2,8 | 9 | 3,3% | 1.7 – 6.2 | 2,3% | no |
|  | *M. europaeum* | 0 | 0,0% | 0,0 – 1,9 | 9 | 3,3% | 1.7 – 6.2 | 3,3% | yes |
|  | *M. branderi* | 0 | 0,0% | 0,0 – 1,9 | 7 | 2,6% | 1.2 – 5.2 | 2,6% | yes |
| **II** | **Fast-growers** | 4 | 2,1% | 0,8 – 5,1 | 5 | 1,8% | 0.7 – 4.2 | -0,2% | no |
| 1 | M. abscessus | 2 | 1,0% | 0,2 – 2,8 | 2 | 0,7% | 0.1 – 2.6 | -0,3% | no |
| 2 | M. fortuitum | 2 | 1,0% | 0,2 – 2,8 | 2 | 0,7% | 0.1 – 2.6 | -0,3% | no |
| 3 | M. stephanolepidis | 0 | 0,0% | 0,0 – 1,9 | 1 | 0,4% | 0.0 – 1.8 | 0,4% | no |

The study analyzed 193 cases of NTM-associated diseases in patients without COVID-19 and 271 cases in patients with COVID-19. It was found that the distribution of mycobacterial species in the two groups differed statistically significantly (chi-square test for homogeneity, χ^2^ ≈ 49.7, p < 0.0001).

A key finding was that COVID-19+ group showed a 28% reduction in MAC prevalence compared to the non-COVID-19 group (94,8% vs 66,8%, p<0,05), mostly associated with reduction in prevalence of *M. avium* (80,3% vs 52,0%, Δ=-28,3%, p<0,05) and *M. chimaera/intracellulare* (12,4% vs 8,5%, Δ=-3,9%, p>0,05) and an increase in prevalence of other NTM. At the same time, an increase in the incidence of rare MAC species was detected: *M. colombiense* (0,5% vs 3,3%, Δ=+2,8%, p<0,05) and *M. celatum* (1,6% vs 3,0%, Δ=+1,4%, p>0,05).

The Mycobacterium kansasii complex was detected in 8.1% of COVID-19+ cases, comprising *M. kansasii* (4.8%) and M. gastri (3.3%), whereas these microorganisms were not detected in the non-COVID-19 group (p < 0.05). A similar pattern was observed for the M. terrae complex, where *M. heckeshornense* was found exclusively in COVID-19+ patients at a frequency of 3.3%.

The *M. simiae* complex demonstrated a marked increase in prevalence, from 0.5% in the the non-COVID-19 group to 7.0% among COVID-19+ patients (p < 0.05). This growth was primarily driven by M. lentiflavum (4.1%) and M. asiaticum (3.0%). A notable rise in the proportion of non-classifiable mycobacterial species was also observed, from 2.6% to 12.9% (p < 0.05), largely attributable to M. europaeum (3.3%) and M. branderi (2.6%).

It is important to note that although a statistical difference was obtained for certain species (e.g., M. gastri and M. asiaticum), which were rare in our study, using the chi-square and Fisher’s exact tests, a comparison of their 95% confidence intervals (CIs) revealed an overlap, likely due to the small sample size.

Furthermore, the differences in the isolation frequency of rapidly growing mycobacteria (M. abscessus, M. fortuitum, M. stephanolepidis) between the COVID-19– and COVID-19+ groups were negligible, at less than 1%.

This study revealed significant alterations in the spectrum of NTM among COVID-19– and COVID-19+ groups of patients with NTM-associated diseases. The analysis of 193 cases in the COVID-19-negative group and 271 cases in the COVID-19-positive group demonstrated that slowly growing mycobacteria remained dominant in both cohorts (97.9% and 98.2%, respectively), albeit with pronounced changes at the species level.

The key finding was a sharp decrease in the frequency of the Mycobacterium avium complex (MAC) in the COVID-19-positive group, from 94.8% to 66.8%. This reduction was primarily driven by a decline in the proportion of M. avium itself, from 80.3% to 52.0%. Concurrently, a statistically significant increase in the prevalence of other mycobacterial species was observed. Notably, the M. kansasii complex (8.1%), the M. terrae complex (3.3%), and the M. simiae complex (7.0%) were detected in the COVID-19-positive group but were virtually absent in the control group. An increase in the frequency of non-classifiable species was also noted, rising from 2.6% to 12.9%.

These findings suggest that COVID-19 impacts the respiratory tract microbial landscape, creating conditions favorable for the colonization and growth of specific NTM species that are considerably less common under normal circumstances. The obtained results necessitate a reevaluation of the diagnostic and therapeutic approaches for mycobacterial infections in COVID-19-positive patients, taking into account the altered spectrum of potential pathogens.

Our findings indicate that SARS-CoV-2 infection may establish specific conditions conducive to the proliferation of certain NTM species, while suppressing others. This may be explained by direct viral effects on the host immune system, as well as indirect consequences, including therapy-induced alterations in the airway microbiota.

Several mechanisms may explain these observations. COVID-19-induced respiratory tract inflammation and immune modulation could create microenvironmental conditions favoring specific NTM species while suppressing others. The cytokine storm associated with severe COVID-19 may alter local immune responses, affecting NTM colonization patterns [28].

Additionally, COVID-19 treatment protocols, including corticosteroids and broad-spectrum antibiotics, may indirectly influence respiratory microbiota composition, potentially affecting NTM species distribution. The prolonged hospitalization often required for severe COVID-19 may also increase exposure to healthcare-associated NTM strains.

Our findings highlight the need to adapt diagnostic and therapeutic strategies for nontuberculous mycobacterial (NTM) disease in post-COVID-19 patients. Updated protocols must include targeted detection of rare mycobacterial species, which often require specific treatment regimens. Furthermore, this study affirms the critical importance of species-level identification for the effective management of NTM infections in the post-COVID era.

The emergence of rare NTM species in post-COVID patients has important clinical implications. Species like M. kansasii, M. heckeshornense, and M. lentiflavum often require specific therapeutic approaches different from standard MAC treatment regimens [19]. Their increased prevalence necessitates enhanced diagnostic capabilities and updated treatment protocols.

The clinical significance of detecting previously uncommon NTM species remains to be fully elucidated. Some may represent colonization rather than active disease, while others could indicate emerging pathogenic potential in immunocompromised hosts [8].

Our study is among the first to provide data on the NTM among patients with NTM-associated diseases who have had COVID-19 in Russia. Despite the statistically significant results obtained, our research has several important limitations. First, the absence of certain mycobacterial species in the nosological structure of NTM-associated diseases in the non-COVID-19 group is noteworthy, particularly the lack of such a significant pathogen as M. kansasii, which, according to the literature, is a frequent cause of NTM-associated diseases in Russia. The detection and identification of NTM is a complex process, and the quality and efficiency of mycobacterial detection directly depend on the laboratory’s equipment and the algorithms used. We cannot rule out that the observed differences may be attributed not only to a prior COVID-19 infection but also to specific features of the laboratory diagnostics used during the initial diagnosis of NTM-associated diseases.

Secondly, as repeatedly highlighted in both Russian and international publications, unlike M. tuberculosis and tuberculosis (TB), NTM and NTM-associated diseases are not subject to mandatory registration. Furthermore, it is possible that some instances of NTM detection were not classified as clinical disease, and vice versa. In our study, this may have led to the under-ascertainment of patients with NTM and, consequently, to the underreporting of NTM-associated diseases cases, irrespective of COVID-19 status. To overcome these limitations, the introduction of mandatory registration for all NTM detections and associated NTM-associated diseases cases, along with standardized, highly efficient algorithms for NTM identification across Russia, is necessary. Despite these limitations, we believe the volume and design of our study are sufficient to support the main conclusions regarding the differences between the two groups and provide valuable information for further research in this area.

## Conclusion

This study reveals significant COVID-19-associated changes in NTM species composition among NTM-associated diseases patients in Russia. The reduced prevalence of MAC and increased detection of rare NTM species highlight the need for adapted diagnostic and therapeutic approaches in the post-COVID era. Species-level identification remains crucial for optimal patient management and antimicrobial stewardship. These findings underscore the complex interactions between viral and bacterial respiratory pathogens and emphasize the importance of continued surveillance to understand evolving NTM epidemiology in the post-pandemic period.

Further research should investigate the mechanistic basis for COVID-19-associated changes in NTM epidemiology. Longitudinal studies tracking NTM species evolution in post-COVID patients would provide valuable insights into temporal relationships and clinical outcomes.

The impact of COVID-19 variants, vaccination status, and specific therapeutic interventions on NTM species distribution warrants investigation. Additionally, standardized diagnostic protocols and mandatory reporting systems for NTM would enhance surveillance and epidemiological understanding.

## Data Availability Statement

The datasets generated and analyzed during the current study are available from the corresponding author upon reasonable request. All relevant data supporting the conclusions of this article are included within the manuscript.

## Author Contributions

PE: Conceptualization, methodology, data analysis, writing—original draft. AB: Data collection, laboratory analysis, writing—review and editing. AK: Laboratory analysis, data interpretation, writing—review and editing. EK: Statistical analysis, data visualization, writing—review and editing. AS: Data collection, methodology, writing—review and editing. IV: Supervision, project administration, writing—review and editing, funding acquisition.

## Acknowledgments

The authors thank the laboratory staff and participating clinical centers across the Russian Federation for their contributions to data collection and analysis. We acknowledge the patients who participated in this study.

## References

1. Narimisa N, Bostanghadiri N, Goodarzi F, Razavi S, Jazi FM. Prevalence of Mycobacterium kansasii in clinical and environmental isolates, a systematic review and meta-analysis. Front Microbiol. 2024;15:1321273.

2. Cowman S, van Ingen J, Griffith DE, Loebinger MR. Non-tuberculous mycobacterial pulmonary disease. Eur Respir J. 2019;54(1):1900250.

3. Griffith D, Aksamit T, Brown-Elliott B, et al. An official ATS/IDSA statement: diagnosis, treatment, and prevention of nontuberculous mycobacterial diseases. Am J Respir Crit Care Med. 2007;175(4):367–416

4. Kim HW, Lee JW, Yoon HS, et al. Restriction of mitochondrial oxidation of glutamine or fatty acids enhances intracellular growth of Mycobacterium abscessus in macrophages. Virulence. 2025;16(1):2454323.

5. Capstick T, Hurst R, Keane J, Musaddaq B. Supporting Patients with Nontuberculous Mycobacterial Pulmonary Disease: Ensuring Best Practice in UK Healthcare Settings. Pharmacy (Basel). 2024;12(4):126.

6. Akutsu T, Tone K, Hasegawa A, et al. Challenges in achieving the guideline-recommended amikacin level for Mycobacterium avium complex pulmonary disease. J Clin Tuberc Other Mycobact Dis. 2025;39:100514.

7. Axson E.L., Bual N., Bloom C.I., Quint J.K. Risk factors and secondary care utilisation in a primary care population with non-tuberculous mycobacterial disease in the UK. Eur. J. Clin. Microbiol. Infect. Dis. 2019;38:117–124. doi: 10.1007/s10096-018-3402-8

8. Opperman C, Steyn J, Matthews MC, Singh S, Ghebrekristos Y, Kerr TJ, Miller M, Esmail A, Cox H, Warren R, Ghielmetti G, Goosen W. Targeted deep sequencing of mycobacteria species from extrapulmonary sites not identified by routine line probe assays: A retrospective laboratory analysis of stored clinical cultures. IJID Reg. 2024 Sep 24;13:100464. doi: 10.1016/j.ijregi.2024.100464. PMID: 39483154; PMCID: PMC11526053.

9. Kim SH, Jhun BW, Jeong BH, Park HY, Kim H, Kwon OJ, Shin SH. The Higher Incidence of COVID-19 in Patients With Non-Tuberculous Mycobacterial Pulmonary Disease: A Single Center Experience in Korea. J Korean Med Sci. 2022 Aug 15;37(32):e250. doi: 10.3346/jkms.2022.37.e250. PMID: 35971764; PMCID: PMC9424694.

10. TB/COVID-19 Global Study Group. Tuberculosis and COVID-19 co-infection: description of the global cohort. Eur Respir J. 2022;59(3):2102538.

11. Пандемия COVID-19 и ее влияние на течение других инфекций на Северо-Западе России / H. A. Беляков, E. B. Боева, O. E. Симакина [и др.] // ВИЧ-инфекция и иммуносупрессии. – 2022. – T. 14, № 1. – C. 7-24. – DOI 10.22328/2077-9828-2022-14-1-7-24. – EDN VXIHJC

12. Малеев, B. B. Некоторые аспекты эволюции инфекционной патологии на современном этапе / B. B. Малеев // Кубанский научный медицинский вестник. – 2020. – T. 27, № 4. – C. 18-26. – DOI 10.25207/1608-6228-2020-27-4-18-26. – EDN MEEMEF.

13. Dahl VN, Mølhave M, Fløe A, van Ingen J, Schön T, Lillebaek T, Andersen AB, Wejse C. Global trends of pulmonary infections with nontuberculous mycobacteria: a systematic review. Int J Infect Dis. 2022 Dec;125:120–131. doi: 10.1016/j.ijid.2022.10.013. Epub 2022 Oct 13. PMID: 36244600.

14. Clinical recommendations tuberculosis in adults, 2024 https://cr.minzdrav.gov.ru/recomend/16_3

15. Daley CL, Iaccarino JM, Lange C, et al. Treatment of Nontuberculous Mycobacterial Pulmonary Disease: An Official ATS/ERS/ESCMID/IDSA Clinical Practice Guideline. Clin Infect Dis. 2020;71(4):e1–e36

16. Микобактериозы легких: сложности диагностики и лечения (обзор литературы) / A. И. Анисимова, M. B. Павлова, Л. И. Арчакова [и др.] // Медицинский альянс. – 2020. – T. 8. – № 1. – C. 25–31.

17. Микобактериозы: современное состояние проблемы/ Зимина B.H.1, Дегтярева C. Ю., Белобородова E.H., Кулабухова E. И., Русакова Л.И., Фесенко O.B.// Клиническая микробиология и антимикробная химиотерапия. – 2017. – T. 19. – № 4. – C. 276–282.

18. Микробиологические и эпидемиологические особенности микобактериозов / И. B. Петров, T. H. Амирова, Л. B. Петрова [и др.] // Эпидемиология и вакцинопрофилактика. – 2020. – T. 19, № 3. – C. 89-94. – DOI 10.31631/2073-3046-2020-19-3-89-94. – EDN GYYQWD.

19. van Tonder AJ, Ellis HC, Churchward CP, Kumar K, Ramadan N, Benson S, Parkhill J, Moffatt MF, Loebinger MR, Cookson WOC. Mycobacterium avium complex genomics and transmission in a London hospital. Eur Respir J. 2023 Apr 20;61(4):2201237. doi: 10.1183/13993003.01237-2022. PMID: 36517182; PMCID: PMC10116071.

20. Masoumi M, Sakhaee F, Vaziri F, Siadat SD, Fateh A. Reactivation of Mycobacterium simiae after the recovery of COVID-19 infection. J Clin Tuberc Other Mycobact Dis. 2021 Aug;24:100257. doi: 10.1016/j.jctube.2021.100257. Epub 2021 Jun 25. PMID: 34222683; PMCID: PMC8233063.

21. Wang X., Ding S., Niu Y. et al. A Comprehensive Review and Update on Epidemiology, Symptomatology, and Management of Nontuberculous Mycobacteria (NTM) // Microbiology Journal. 2022. URL: https://microbiologyjournal.org/a-comprehensive-review-and-update-on-epidemiology-symptomatology-and-management-of-nontuberculous-mycobacteria-ntm/.

22. Rhoads D.D., Novak-Weekley S.M. Non-tuberculous mycobacteria pulmonary disease: A review of pathogenesis, diagnosis and treatment // Pulmonary Journal. 2022. URL: https://pmc.ncbi.nlm.nih.gov/articles/PMC9394508/.

23. Oliveira G.P., Oliveira A.C., Silva L.S., et al. Rates of infection with other pathogens after a positive COVID-19 test // The Lancet Infectious Diseases. 2025. Vol. 25, No. 3, pp. 310–319. DOI: 10.1016/S1473-3099(24)00012-8. URL: https://www.thelancet.com/journals/laninf/article/PIIS1473-3099(24)00012-8/fulltext.

24. Otero A., Smith J., Garcia M., et al. Analysis reveals global post-COVID surge in infectious diseases // BMJ. 2024. Vol. 375, e067890. DOI: 10.1136/bmj-2024-067890. URL: https://www.bmj.com/content/375/bmj-2024-067890.

25. Ashraf A.M. et al. COVID-19 story: Entry and immune response // Vacunas. 2024. URL: https://www.sciencedirect.com/science/article/abs/pii/S1576988724000992.

26. Cocca S. et al. COVID-19: from immune response to clinical intervention // Precision Clinical Medicine, 2024. URL: https://academic.oup.com/pcm/article/7/3/pbae015/7716697.

27. Romagnoli A., Iantomasi T., Pagliuca C., et al. Cellular and Molecular Immune Responses during Mycobacterium avium complex Infection: A Review // International Journal of Molecular Sciences. 2021. Vol. 22, No. 18, Article 9971. DOI: 10.3390/ijms22189971. URL: https://www.mdpi.com/1422-0067/22/18/9971.

28. Karki R, Kanneganti TD. The ‘cytokine storm’: molecular mechanisms and therapeutic prospects. Trends Immunol. 2021;42(8):681–705. doi:10.1016/j.it.2021.06.001

